# Feasibility, acceptability and preliminary effectiveness of the real-world implementation of informal caregiver support program via text message

**DOI:** 10.1101/2025.09.19.25336094

**Authors:** Jaime Perales-Puchalt, Christina Baker, Binod Wagle, Melissa Godar, Sayra Nieto Gomez, Hayden Johnson, Betty Drees, Mónica Fracachán-Cabrera, Mariana Ramírez-Mantilla

## Abstract

Few dementia caregiver support programs have been tested in real-world settings. We tested the feasibility and preliminary effectiveness (e.g., preparedness for caregiving) of the first short message service (SMS) texting program to support informal caregivers of people with dementia. We analyzed data from 147 caregivers of people with dementia participating in a service program. This program was a remote, asynchronous, and bidirectional texting program focused on dementia education, skill-building, and community resources that lasted six months. We measured outcomes via surveys and metrics of intervention usage over six months. Two caregivers experienced technical issues during the program, 12.9% unsubscribed, and 68.8% read most texts thoroughly. Most caregivers (64.3-75.9%) reported high or very high levels of acceptability. Levels of preparedness for caregiving and unmet needs improved from pre- to post-program testing. This real-world implementation of a texting caregiver support program led to improvements in caregiver outcomes.

## Introduction

Informal caregiving for people with dementia (PWD) is highly prevalent and leads to important negative health outcomes. Nearly 12 million people in the US provide unpaid care for a family member or friend with dementia (Alzheimer’s Association, 2024). Negative mental health outcomes of caregiving primarily involve distress, which is apparent in both self-reports and cortisol levels (Corrêa et al., 2015). Caregivers of PWD are also more likely than non-caregivers and caregivers of other populations to have depression (Atteih et al., 2015; Cuijpers, 2005; Sallim et al., 2015; Thunyadee et al., 2015) and anxiety disorders (Atteih et al., 2015; Sallim et al., 2015).

Several efficacious caregiver support interventions exist, including psychoeducational, cognitive behavioral therapy, counseling, support groups, PWD training, respite and multicomponent interventions (Walter & Pinquart, 2020). However, many of the existing interventions rely heavily on workforce to deliver lengthy programs using rigid delivery modalities (mostly live and in-person) which challenges their implementation. In addition, most caregiver interventions have been designed for urban, non-Latino Whites and results do not generalize to underserved groups (Gilmore-Bykovskyi et al., 2018). Underserved groups have a higher risk of developing dementia, provide more intense and longer caregiving, experience higher levels of caregiver distress, depression, and burden, and are less likely to attend caregiver support services despite their interest in receiving support (Alzheimer’s Association, 2024; Hudomiet et al., 2022; Weden et al., 2018). There is a need for easily implementable caregiver support solutions that do not leave anyone behind.

A potential solution to the barriers above-mentioned is the delivery of content tailored to different cultural and contextual groups via Short Message Service (SMS) texting. SMS texting is highly accessible because nearly all adults nowadays have access to cell phones with SMS texting capabilities, and text messages are remote and asynchronous, meaning that time and space are not barriers to the delivery of caregiver support (Duggan, 2013; Pew Research Center, 2021). We had originally developed and shown the promising feasibility and preliminary efficacy outcomes of the first SMS texting program to deliver caregiver support to Latino informal caregivers, CuidaTEXT (Perales-Puchalt, Acosta-Rullan, et al., 2021; Perales-Puchalt et al., 2022). In 2022, we were approached to adapt CuidaTEXT to the broader community in a Midwest US region and implement it as a service. In this manuscript, we report the results from the implementation of this program (CareTEXT) in our region. We hypothesized that caregivers’ levels of strain would decline following their use of CareTEXT.

## Methods

This is a secondary analysis of a pre-post-intervention assessment of a remote caregiver support quality improvement program that took place from August 2022 to June 2024. One service/research institution administered CareTEXT to a convenience sample of caregivers in a five-county service region in the Midwest US. Recruitment sources included community events, social media, medical records and neurology clinic visits, research registries, word of mouth, and referrals from partner organizations and clinics. Caregivers’ eligibility criteria for the program included speaking English or Spanish, being at least 18 years old, the PWD living in one of the 5 eligible counties, owning a cell phone with a flat fee for texting, being able to read and write texts, emotionally or physically taking care of someone with a clinical or research diagnosis of dementia, and being willing to participate in the program. Due to the service-oriented and non-research priority of the program, we decided to stop asking the full survey and only ask a basic satisfaction question to caregivers halfway through the program period. For this reason, we include data from caregivers who were administered the full baseline survey only in the current analytic sample (147 out of the 301 caregivers that used the program). The program allowed more than one caregiver per person with dementia but did not identify those caregivers who cared for the same PWD. The University of Kansas Medical Center deemed this project to be a Quality Improvement project. All caregivers gave oral informed consent to participate in the program.

### Intervention

CareTEXT is a six-month bilingual SMS remote and asynchronous program where caregivers receive scheduled messages and can also request on-demand assistance by texting. This intervention, and its development process, have been extensively detailed previously (Perales-Puchalt, Acosta-Rullan, et al., 2021). *CareTEXT* entails sending 1-3 automated daily messages covering various aspects such as logistics, dementia education, self-care, social support, end-of-life care, managing dementia-related behaviors, problem-solving strategies, and community resources. Additionally, participants can text keyword-based queries for immediate assistance on the above-mentioned topics and engage in live chat sessions with the coach for further guidance upon request. The original program (CuidaTEXT) was developed for Latinos.

However, we adapted the program to increase its acceptability among other frequent community groups in the region, namely Black Americans and [mostly White] rural Americans. Work to conduct the adaptations followed Cultural Accommodation Model principles of co-creation with the target communities (Burrow-Sanchez et al., 2011). For this reason, we held five, one-hour videocall meetings with five representatives of each of the two working groups to go over text messages and report whether they liked it the way it was or whether it required an adaptation.

Adaptation suggestions were documented in a word file and were later discussed by the research team, who created a new version and presented it to the working groups to generate a final version. Adaptations included reducing the number of names in Spanish and increasing names typical of other cultures, changing some expression and terms, adding new contexts (e.g., referencing the Deep South and the Church), and references and links to culturally-relevant lifestyle behaviors such as line dancing and soul foods.

### Assessment

The research team collected information from three sources: baseline survey, six-month follow-up survey, and metrics of text message interactions. We collected sociodemographic and care-related information from the caregivers at baseline.

Outcomes included feasibility, acceptability, and preliminary efficacy:

<u>Feasibility outcomes</u> were similar to those reported in our previous study (Perales-Puchalt et al.). Outcomes included the feasibility of intervention enrollment (percentage of participants who were able to opt into CareTEXT), retention rate (completing follow-up survey), assessment (completing preliminary efficacy outcomes), intervention delivery (technical issues), and intervention engagement (usage of texting features, unsubscribing, and reports of attention paid to CareTEXT).

<u>Acceptability outcomes</u> were also like the ones used in our previous study (Perales-Puchalt et al.). We collected all of them in the six-month follow-up survey. These include four questions: The first three questions asked to what extent CareTEXT had helped understand dementia more, and helped care for oneself and the PWD. These questions had four response options ranging from not at all to a lot. A fourth question asked about the overall satisfaction with CareTEXT, with four response options ranging from not at all to extremely.

<u>Preliminary efficacy outcomes</u> included three scales about the caregiver. All scales have shown good psychometric properties previously in their original publications, except for the unmet needs checklist, which the research team developed:

<u>Depressive symptoms (Center for Epidemiologic Studies Depression Scale [CES-D-10] (Cheng & Chan, 2005))</u>: This is a 10-item, self-report rating scale that measures characteristic symptoms of depression in the past week (e.g., depression, loneliness). Each item is rated on a 4-point scale, from 0 (Rarely or None of the Time) to 3 (Most or All of the Time) with positively worded items (items 5 and 8) reverse scored. Items yield summary scores that range from 0 to 30, with higher scores indicating higher depression severity.

<u>Caregiver strain (Modified Caregiver Strain Index [CSI] (Thornton & Travis, 2003)):</u> The CSI is a 13-item screener that measures caregiver strain. For each of the items (e.g., caregiving is inconvenient), the caregiver can respond either No (0), Yes Sometimes (1), or Yes on a Regular Basis (2). The total score is the sum of all item scores. Higher scores indicate higher strain.

<u>Caregiving competence (Preparedness for Caregiving Scale [PCS] (Carter et al., 1998; Gutierrez-Baena & Romero-Grimaldi, 2021)):</u> The PCS is a self-rated instrument that consists of eight items that ask caregivers how well prepared they believe they are for multiple domains of caregiving (e.g., caring for care recipient’s physical needs). Responses are rated on a 5-point Likert scale with scores ranging from 0 (Not at All Prepared) to 4 (Very Well Prepared). The scale is scored by calculating the mean of all items answered, with a total score range of 0 to 4. The higher the score the more prepared the caregiver feels for caregiving.

<u>Unmet needs checklist</u>: This checklist was created by the contracting institution as a way to inform the resources caregivers would need to get help with. This is a seven-item checklist for caregivers that requires 0 (No) or 1 (Yes) answers. The checklist states “In addition to the services you are already receiving, do you need any more services?”. The options include 1) connection to information and resources, 2) respite, 3) legal advice/assistance, 4) emotional/social support, 5) additional training, 6) home repairs or modifications to make caregiving tasks safer, and 7) other. A total score is obtained by summing all seven items. The higher the score, the higher the number of unmet needs the caregiver reports having. The unmet needs checklist’s internal consistency is not appropriate because the checklist does not measure overlapping domains. The scale’s concurrent validity using Spearman correlation with the CSI was 0.286; p<0.01.

### Analysis

We used descriptive statistics to summarize the baseline characteristics, and to report feasibility and acceptability outcomes. To assess preliminary efficacy, we conducted paired-samples t-tests to evaluate changes from baseline to follow-up. For these t-tests, we also report effect sizes using Cohen’s d’s thresholds of small (0.2), medium (0.5), and large (0.8) (Cohen, 2013). We performed all analyses using SPSS Version 22 (IBM Corp., 2023). The significance level was set at p < 0.05.

## Results

Table 1 summarizes the baseline characteristics of caregivers. The sample averaged 63.6 years of age (SD 11.1; min 29; max 85) and 15.4 years of education (SD 2.5) and most reported being women (84.4%; n=124), White (79.6%; n=117), urban-living individuals (84.4%; n=124), that were married or live with a partner (76.7%; n=112), had no difficulties in paying for basic needs (12.4%; n=18), were the partner of the PWD (51.0%; n=75), and their PWD had either an Alzheimer’s disease (37.4%; n=55) or an unknown dementia diagnosis (31.3%; n=46). Completion of the follow-up assessment tended to be associated with being older, male, non-White, more education, married or living with a partner, having no difficulty paying for basic needs, being a partner of the PWD, and having a known dementia diagnosis.

**Table 1.** Baseline characteristics of the caregivers being served in the CareTEXT program for the total sample, those who completed and the follow-up survey (completers), and those who did not (non-completers)

| Caregivers | Total<br>(n=147) | Completers<br>(n=113) | Non-completers<br>(n=34) |
| --- | --- | --- | --- |
| Age in years, mean (SD) | 63.6 (11.1) | 64.9 (10.4) | 59.4 (12.6) |
| Women, % (n) | 84.4% (124) | 82.3% (93) | 91.2% (31) |
| Latino ethnicity, % (n) | 8.2% (12) | 8.8% (10) | 5.9% (2) |
| Race, % (n) |  |  |  |
| White, % (n) | 79.6% (117) | 78.8% (89) | 82.4% (28) |
| Black, % (n) | 14.3% (21) | 15.0% (17) | 11.8% (4) |
| Other, % (n) | 6.1% (9) | 6.2% (7) | 5.8% (2) |
| Rural residence, % (n) | 15.6% (23) | 15.0% (17) | 17.6% (6) |
| Years of education, m (SD) | 15.4 (2.5) | 15.5 (2.3) | 14.8 (3.0) |
| Without medical insurance, % (n) | 2.7% (4) | 2.7% (3) | 3.0% (1) |
| Married or have a partner, % (n) | 76.7% (112) | 81.4% (92) | 73.6% (25) |
| Difficult or very difficult to pay for basic needs, % (n) | 12.4% (18) | 9.7% (11) | 21.9% (7) |
| Relation to PWD |  |  |  |
| Partner, % (n) | 51.0% (75) | 57.5% (65) | 29.4% (10) |
| Children, % (n) | 29.9% (44) | 24.8% (28) | 47.1% (16) |
| Other, % (n) | 19.5% (28) | 17.7% (20) | 23.5% (8) |
| Dementia type of the PWD |  |  |  |
| Alzheimer's disease, % (n) | 37.4% (55) | 38.9% (44) | 32.4% (11) |
| Unknown, % (n) | 31.3% (46) | 26.5% (30) | 47.1% (16) |
| Mixed dementia, % (n) | 9.5% (14) | 12.4% (14) | 2.9% (1) |
| Lewy bodies/Parkinson's dementia, % (n) | 10.2% (15) | 9.7% (11) | 8.8% (3) |
| Other dementia, % (n) | 11.6% (17) | 12.4% (14) | 8.8% (3) |

Feasibility and acceptability findings can be seen on Table 2 and Figure 1, respectively. Out of the 147 caregivers who completed the full baseline survey, all were able to opt into CareTEXT, 113 (76.9%) completed at least the acceptability section of the follow-up survey and 100 (68.0%) completed the full survey, since their PWD was still alive. Ony two caregivers reported having [small] technical issues with CareTEXT, and these were related to messages not coming through at a certain point in time. Most caregivers (68.8%; n=77) read most messages thoroughly, 12.9% (n=19) unsubscribed from the program before it ended, and 81.0% (n=119) sent at least one text message to the program. Most participants perceived CareTEXT helped them very or extremely understand dementia more (75.9%); and care for themselves (66.3%), care for their PWD (70.6%), and most were very or extremely satisfied with CareTEXT (73.2%).

**Table 2.** Feasibility of *CareTEXT*

| Feasibility |  |  |
| --- | --- | --- |
| Intervention enrollment | Percentage of respondents able to opt into *CareTEXT*<sup>a</sup> | 100%<br>(n=147) |
| Retention rate | Percentage of enrolled respondents who completed the follow-up survey<sup>a</sup> | 76.9%<br>(n=113) |
| Assessment feasibility | Percentage of enrolled respondents who completed the preliminary efficacy outcomes<sup>a</sup> | 68.0%<br>(n=100) |
| Intervention delivery | Percentage of respondents who experienced technical issues: First message took a few days to come through, 2) some issues with reception of messages<sup>b</sup> | 1.4% (n=2) |
| Intervention engagement | Read through most messages thoroughly<sup>b</sup> | 68.8% (77) |
|  | Took a short look at messages most times<sup>b</sup> | 30.4% (34) |
|  | Did not read messages most times<sup>b</sup> | 0.9% (1) |
|  | Percentage of respondents who unsubscribed from *CareTEXT*<sup>a</sup> | 12.9% (19) |
|  | Percentage of respondents who sent 0 text messages (no interaction)<sup>a</sup> | 19.0%<br>(n=28) |
|  | Percentage of respondents who sent 1-9 text messages (low interaction)<sup>a</sup> | 38.1%<br>(n=56) |
|  | Percentage of respondents who sent 10-49 text messages (intermediate interaction)<sup>a</sup> | 32.7%<br>(n=48) |
|  | Percentage of respondents who sent 50-99 text messages (high interaction)<sup>a</sup> | 7.5% (n=11) |
|  | Percentage of respondents who sent more than 100 text messages (very high interaction)<sup>a</sup> | 2.7% (n=4) |

a: calculated for all caregiver participants (n=100); b: calculated for caregiver completers (n=113)

**Figure 1.**
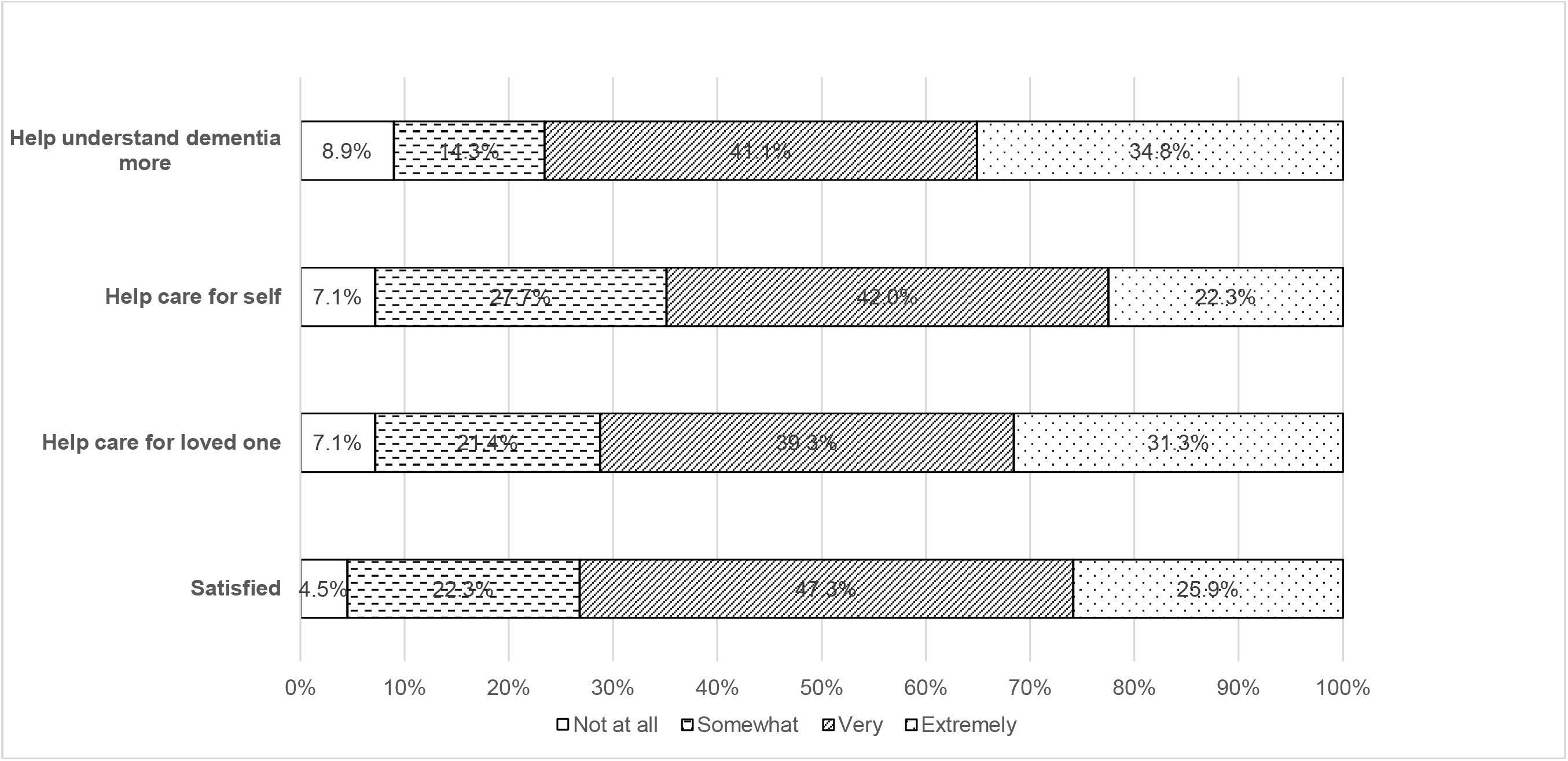
Acceptability of CareTEXT (n=113)

Preliminary effectiveness can be seen in table 3. Participation in CareTEXT was not followed by an improvement in caregiver depression as rated with the CES-D-10 or strain as rated with the CSI. However, two outcomes improved after participating in CareTEXT. Outcomes that improved included preparedness for caregiving as measured by the PCS, which increased from 2.3 to 2.5 (95% confidence interval of mean difference [95% CI]=-0.34; -0.09; Cohen’s d=0.33), and unmet needs, which decreased from 1.7 to 0.9 (95% CI of mean difference= 0.51; 1.05; Cohen’s d=0.56).

**Table 3.** Preliminary effectiveness outcomes comparing pre- and post-intervention scores

| Caregiver | n | M Baseline | M Follow up | M difference | SD of M difference | 95% CI Lower | 95% CI Upper | Cohen's d |
| --- | --- | --- | --- | --- | --- | --- | --- | --- |
| CES-D-10 depression (0-30) | 95 | 9.3 | 8.6 | 0.69 | 5.00 | -0.32 | 1.71 | 0.14 |
| CSI Strain (0-24) | 97 | 10.5 | 11.3 | -0.77 | 4.53 | -1.68 | 0.15 | 0.17 |
| PCS Preparedness (0-4) | 100 | 2.3 | 2.5 | -0.21 | 0.65 | -0.34 | -0.09 | 0.33 |
| Unmet needs (0-7) | 100 | 1.7 | 0.9 | 0.8 | 1.38 | 0.51 | 1.05 | 0.56 |

## Discussion

In this study, we described the feasibility, acceptability and preliminary effectiveness findings from a real-world implementation of CareTEXT among informal caregivers. This is the first time an SMS texting program for dementia caregiving support has been implemented in the real world. Results suggest that CareTEXT can be implemented feasibly and satisfactorily in the real world among people of different ethnic, racial and urban/rural setting backgrounds. Rather than decreasing depressive symptoms or strain, CareTEXT might be more likely to increase caregiver preparedness for caregiving and reduce unmet needs in this population.

Our findings add to the scarce literature of dementia caregiver support real-world program implementation. Only 15 dementia caregiver support programs have been tested in the real world (Hodgson & Gitlin, 2021; Hwang et al., 2024). Some of these include well-established programs such as the Resources for Enhancing Alzheimer’s Caregiver Health II, Reducing Disability in Alzheimer’s Disease, and Savvy Caregiver (Burgio et al., 2009; Griffiths et al., 2015; Perales-Puchalt, Barton, et al., 2021). CareTEXT reached high levels of satisfaction in a diverse population of dementia caregivers. These positive findings align with recommendations to tailor programs to groups from multiple ethnic, racial and rural backgrounds for caregiver support implementations to succeed (Hodgson & Gitlin, 2021). To our knowledge, this is the only dementia caregiver support program that relies largely on SMS texting. SMS texting is readily available to most caregivers given that most adults own a cell phone with SMS texting capabilities, and SMS texting asynchronous nature allows caregivers to access the content of the program at anytime and anywhere (Duggan, 2013; Pew Research Center, 2021). This level of accessibility might explain the high intervention enrollment and engagement in CareTEXT.

Some feasibility and acceptability outcomes are similar and different to previous versions of the current SMS texting program and SMS texting programs for other conditions. CareTEXT caregivers had a lower engagement with text messages than previous versions. For example, previously, 25% of caregivers texted 100 messages or more during the six months of the program. However, in the current study, the percentage was 2.7%. Similarly, 86% reported reading most texts thoroughly in the previous study vs 69% of our current sample. A cause for the higher engagement in the previous study might be the inclusion of a largely non-Latino and older sample. Latinos and younger generations engage more with SMS texting than other populations (Duggan, 2013), and Latinos’ baseline dementia knowledge is lower (Connell et al., 2007). While unsubscription rates were higher than our previous study (13% vs 0%), they were still lower than texting programs for other conditions that were not co-developed with the target audience (e.g., 34% within 4 weeks) (Abroms et al., 2015). These results speak to the importance of tailoring to different populations by including them in the design. Real-world implementation of caregiver support interventions that relied on in-person meetings has also achieved lower program-engagement. For example, a local adaptation of Reducuing Disability in Alzheimer’s Disease achieved 63% completion of nine out of the 12 modules of the program (Perales-Puchalt, Barton, et al., 2021). These findings point to the importance of available remote asynchronous modalities for the delivery of programs.

CareTEXT achieved improved preliminary effectiveness outcomes. These outcomes included preparedness and unmet needs. CareTEXT increased preparedness for caregiving, which is in line with CareTEXT’s original study with the Latino population (Perales-Puchalt et al., 2022).

Improving preparedness for caregiving is important because it has been described as a target for dementia caregiving support interventions, given its association with improvement with multiple other caregiver and care recipient mental health and safety outcomes (Hancock et al., 2022). The lack of an effect on caregiver depressive symptoms and strain is not in line with our previous version of CareTEXT in the Latino population but aligns with other caregiver interventions. For example, telephone-based support groups and a video-phone psychosocial intervention led to no improvements in depressive symptoms six months after baseline (Czaja et al., 2013; Martindale-Adams et al., 2013). Our study resulted in the same percentage of reduction of unmet needs as a real-world dyad home-based behavior management and exercise program (47%) (Perales-Puchalt, Barton, et al., 2021). Another real-world implementation of the same home-based program achieved similar results, including a 35% reduction in unmet needs (Menne et al., 2014).

The current study has limitations. This was a quality improvement research project of a service program, which might reduce the generalizability of findings due to its local nature and non-research priority focus. Similarly, we did not include a control group and therefore cannot attribute causality of improvements in outcomes to participating in the program. We did not report fidelity, which is important for the implementation of programs in the real-world (Hwang et al., 2024). Since the implementation relied on the same team as the research, we also did not analyze other implementation science outcomes such as the adoption or reach of interventionists. However, our program implementation ended along with the end of funding, which indicates a reliance on funding for sustainability, mostly to fund the time for enrollment and coach support. There was a 23% of non-completion of follow-up surveys. These individuals could have had more undesirable feedback that was not assessed than those who completed the follow-up surveys.

This study has implications for public health and future research. The high feasibility, acceptability outcomes and improvement in preliminary effectiveness outcomes warrant future funding for the implementation of this program to serve dementia caregivers among diverse populations. However, to do so, it is crucial either to identify funding mechanisms, or to develop fully automatic versions of this intervention to offset the cost of coaches. A previous fully powered efficacy study is more pressing, as this has not yet been established. Once efficacy has been established, studies could also assess the population impact of this vs in-person interventions, as this would consider not just effectiveness, but also reach. Future studies should analyze the fidelity of the intervention. While this analysis is outside the scope of the current study, we have tracked all interactions between the caregivers and CareTEXT automatic and coach-based messages. Future studies should also assess barriers and facilitators of implementing CareTEXT in other settings such as the dementia associations, aging agencies and health care settings.

## Conclusion

This work contributes to the scarce literature on real-world implementation of caregiver support interventions, by reporting results from the first SMS texting caregiver support intervention. Findings indicate that, as soon as CareTEXT shows efficacy, it can be implemented with feasibility, acceptability and preliminary effectiveness. Future research is warranted to better understand barriers and facilitators to implement this program in other settings and to maximize sustainability.

## Data Availability

The data that support the findings of this study are not publicly available due to institutional restrictions and patient privacy considerations. Requests for de-identified data may be considered by the authors on a case-by-case basis.

## Acknowledgements

JPP thanks the national and local organizations that have partnered with him to conduct present and past research since 2015. Especially the Mid-America Regional Council, James Stowe and Sarah Albin for letting us know about the funds and guiding us through the bureaucracy. The research team thanks the Community Advisory Board who helped adapt the program to diverse populations and all families included in all stages of this project as well as anyone who has contributed directly and indirectly to it. We also thank all platforms and organizations which have shared the opportunity to participate in this project with the people they serve. The ideas and opinions expressed herein are those of the authors alone, and endorsement by the authors’ institutions or the funding agency is not intended and should not be inferred.

